# Progressive dysmorphia of retinal pigment epithelium in age related macular degeneration revealed by fluorescence lifetime imaging

**DOI:** 10.1101/2021.04.14.21255463

**Authors:** Martin Hammer, Juliane Jakob-Girbig, Linda Schwanengel, Christine A. Curcio, Somar Hasan, Daniel Meller, Rowena Schultz

## Abstract

**Purpose:** To observe changes of the retinal pigment epithelium (RPE) on the transition from dysmorphia to atrophy in age related macular degeneration (AMD) by fluorescence lifetime imaging ophthalmoscopy (FLIO).

**Methods:** Multimodal imaging including color fundus photography (CFP), optical coherence tomography (OCT), fundus autofluorescence (FAF) imaging, and FLIO was performed in 40 eyes of 37 patients with intermediate AMD and no evidence for geographic atrophy or macular neovascularization) (mean age: 74.2±7.0 years). Twenty-three eyes were followed for 28.3±18.3 months. Seven eyes had a second follow up after 46.6±9.0 months. Thickened RPE on OCT, hyperpigmentation on CFP, and migrated RPE, seen as hyperreflective foci (HRF) on OCT, were identified. Fluorescence lifetimes in two spectral channels (SSC: 500-560 nm, LSC: 560-720 nm) as well as emission spectrum intensity ratio (ESIR) of the lesions were measured by FLIO.

**Results:** As hyperpigmented areas form and RPE migrates into the retina, FAF lifetimes lengthen and ESRI of RPE cells increase. Thickened RPE showed lifetimes of 256±49 ps (SSC) and 336±35 ps (LSC) and an ESIR of 0.552±0.079. For hyperpigmentation, these values were 317±68 ps (p<0.001), 377±56 ps (p<0.001), and 0.609±0.081 (p=0.001), respectively, and for HRF 337±79 ps (p<0.001), 414±50 ps (p<0.001), and 0.654±0.075 (p<0.001).

**Conclusions:** In the process of RPE degeneration, comprising different steps of dysmorphia, hyperpigmentation, and migration, lengthening of FAF lifetimes and a hypsochromic shift of emission spectra can be observed by FLIO. Thus, FLIO might provide early biomarkers for AMD progression and contribute to our understanding of RPE pathology.

## Introduction

Although age related macular degeneration (AMD) is a multifactorial disease, the end stage of non-neovascular AMD includes atrophy of the retinal pigment epithelium (RPE). This is seen as geographic atrophy (GA) in color fundus photography (CFP) and is incorporated into complete RPE and outer retinal atrophy (cRORA) in optical coherence tomography (OCT).^1^ Consequently, RPE alteration, seen in CFP or OCT, is a risk factor for disease progression.^2-5^ Histopathology as well as clinical imaging revealed multiple morphologically distinct pathways to atrophy, mainly intraretinal migration and shedding of organelles into underlying basal laminar deposits (BLamD).^6, 7^ The best studied pathway involves RPE cells detaching from the RPE basal lamina (BL) and subsequently migrating into the retina, where they appear as hyperreflective foci (HRF) in OCT.^5, 8-10^ In a post-hoc analysis of a standardized imaging dataset from a prospective clinical trial, HRF were found to confer risk for RPE atrophy with a 2-year odds ratio of 5.2.^11^

Due to its accumulation of lipofuscin and melanolipofuscin, RPE contributes the strongest fluorescence emission in fundus autofluorescence imaging (FAF).^12-17^ High resolution microscopy revealed distinct autofluorescent organelles^18^ and spectrally resolved microscopy uncovered emission spectra specific for RPE fluorophores and for sub-RPE tissues.^19^ Hyperautofluorescence in FAF was studied as a marker of AMD progression and GA growth clinically^20-24^ as well as in histopathology.^25^ Furthermore, hyperpigmentation in CFP is associated with hyperautofluorescence in FAF.^16, 17^

Fluorescence lifetime imaging ophthalmoscopy (FLIO) is a technique extending FAF imaging to the measurement of fluorescence lifetimes.^26-28^ Measuring the average time (in the order of picoseconds) a fluorescent molecule remains in an excited electronic state after excitation by a short laser pulse, fluorescence lifetime imaging characterizes the molecule and its embedding matrix. In clinical FLIO imaging, however, a variety of fluorophores can contribute to the signal. As it is impossible to resolve all molecules from the fluorescence decay signal, these measurements characterize the overall state of tissue.^29^ A general prolongation of FAF lifetimes ^30, 31^ as well as further prolongation with disease progression was found in AMD.^32^ Specifically, hyperpigmented areas in CFP exhibited longer FAF lifetimes and shorter emission wavelengths than the surrounding less-affected areas in the fundus.^33^

In the current retrospective study, we used multimodal imaging to investigate the natural history of RPE dysmorphia as defined in CFP and OCT, with the expectation that these cells are abnormal and thus would exhibit distinctive signals in FAF and FLIO. In this study, activated RPE atop drusenoid RPE detachment (PED), hyperpigmentation, and migrated RPE were examined separately in order to observe FAF lifetime changes and identify AMD progression markers seen in FLIO.

## Methods

### Subjects and Procedures

In this retrospective study, patients with non-neovascular AMD seen at the University Hospital Jena, Department of Ophthalmology between April 2014 and January 2021 were included. Exclusion criteria were the presence of GA or macular neovascularization at the baseline visit, other retinal pathologies such as diabetic retinopathy, glaucoma, hypertensive retinopathy, retinal vascular occlusion, central serous chorioretinopathy, macular telangiectasia 2, Stargardt disease, retinitis pigmentosa, macular holes, and retinal detachment. Patients were also excluded in case of mature cataract preventing high quality fundus imaging.

The study was approved by the ethics committee of the University Hospital Jena and adhered to the tenets of the declaration of Helsinki. All participants gave written informed consent prior to study inclusion and underwent a full ophthalmologic examination including best corrected visual acuity, OCT (Cirrus-OCT, Carl-Zeiss Meditec AG, Jena, Germany) and CFP (Visucam, Carl-Zeiss Meditec AG, Jena, Germany). Pupils were dilated using tropicamide (Mydriaticum Stulln, Pharma Stulln GmbH, Nabburg, Germany) and phenylephrine-hydrochloride (Neosynephrin-POS 5%, Ursapharm GmbH, Saarbrucken, Germany). After pupil dilation, patients underwent FLIO imaging, OCT, and CFP. No sodium fluorescein was administered topically or intra-venously prior to FLIO investigation.

### FLIO Imaging and Data Analysis

Basic principles and laser safety of FLIO are described elsewhere.^34-36^ FLIO image capture is based on a picosecond laser diode coupled with a laser scanning ophthalmoscope (Spectralis, Heidelberg Engineering, Heidelberg, Germany), exciting retinal autofluorescence at 473 nm with a repetition rate of 80 Mhz. Fluorescence photons were detected by time-correlated single photon counting (SPC-150, Becker&Hickl GmbH, Berlin, Germany) in short-wavelength (SSC: 498-560 nm) and long-wavelength (LSC: 560-720 nm) spectral channel. FLIO provides 30° field images with a frame rate of 9 frames per second and a resolution of 256 x 256 pixels. Photon histograms over time, describing the autofluorescence decay, were least-square fitted with a series of three exponential functions using the software SPCImage 6.0 (Becker&Hickl GmbH, Berlin, Germany). The amplitude-weighted mean decay time τ_m_ is called FAF lifetime and used for subsequent analysis. The resulting image is color-coded, depicting short lifetimes in red and long lifetimes in blue color. In addition, the ratio of photon counts (autofluorescence intensity) in SSC and LSC was calculated per pixel, which is denoted as the emission spectrum intensity ratio (ESIR).

The software FLIMX, which is documented and freely available for download under an open-source BSD license (http://www.flimx.de),^37^ was used for the manual selection of lesions as regions of interest (ROI). Mean FAF lifetimes and ESIR per pixel were averaged over all pixels of the ROI as well as the environment of the ROI, a 105-µm-wide area which was 35 µm distant to the ROI.

From previous literature, several imaging indicators of dysmorphic RPE and its underlying basal lamina (BL) can be assembled for use in reference to FLIO. The RPE and the RPE-BL are best thought of as separate from an underlying 3-layer Bruch’s membrane^38, 39^. In OCT, a thickening of RPE-BL band can be observed especially over drusen and drusenoid PED ^40^. Thickening precedes the appearance of HRF directly above in the overlying retina but thickening may or may not still be present if and when HRF appears. In non-neovascular AMD, HRF have been directly correlated with nucleated cells containing RPE organelles at the same qualitative abundance as cells in the intact RPE layer^6, 40^. Hyperpigmentation in color fundus photography is a non-specific term that encompasses several possibilities, including proliferation (cell division) of RPE cells, rounding or stacking of individual RPE cells that increase pathlength of light through light-absorbing melanin protein, and increased concentration of melanin-containing organelles (melanosomes and melanolipofuscin).^41^

Two reviewers (JJG and MH), masked to the lifetimes, marked hyperautofluorescent areas in FAF (SSC or LSC) and graded them according to OCT and CFP features: OCT was checked for the presence of drusen or drusenoid PED (smallest diameter > 350 µm^42^), thickening of the RPE-BL band, and the presence or absence of HRF anterior to the ellipsoid zone.^43^ CFP was checked for the presence or absence of hyperpigmentation. This way, we found three lesion entities consistent with the cellular-level model described above: (i) a thickened RPE-BL band atop drusenoid PED with no hyperpigmentation in CFP, (ii) hyperpigmentation associated with thickened RPE-BL but no HRF, and (iii) HRF independent from the thickness of the underlying RPE-BL.

Lesions were excluded if the reviewers graded differently. Areas that were not clearly thickened in OCT and hyperpigmented in CFP were excluded in order to exclude hyperautofluorescent drusen from the analysis. In addition, areas with incipient atrophy were excluded. Lesions in the fovea were excluded as well because of FLIO signal from macular xanthophyll pigment. Acquired vitelliform lesions were also excluded. Grading was done independently for each patient visit.

### Statistics

FAF lifetimes and ESIR of lesions and their environment were compared by paired T-test. Data for groups of lesions (thickened RPE-BL, hyperpigmentation, and HRF) was compared using ANOVA with post-hoc Bonferroni test compensating for multiple testing. For this test, each lesion entity was included at baseline. The correlation of the follow up time with the changes of lifetimes and ESIR from baseline to follow-up was tested.

## Results

Forty eyes of 37 patients (mean age: 74.2±7.0 years) were included. Thirty eyes of 25 patients had a follow up of 28.3±18.3 months. Twelve of these eyes had a second follow up (mean follow up time 46.6±9.0 months after baseline). Study patient demographics and lesion fate are given in Table 1. At baseline, we found 58 areas of thickened RPE atop PED on OCT, 38 areas of hyperpigmentation on CFP without PED on OCT, and 126 HRF. We observed the disappearance of 9 PEDs (6 at first and 3 at second follow up) and 15 HRF at first follow up. Four spots of hyperpigmentation turned to HRF at follow up. Over follow up we found 11 newly appearing PEDs, 7 new spots of hyperpigmentation (3 over drusen and 4 over PED), and 28 new HRF (6 over drusen and 22 over recent or collapsed PED). At first follow up, five out of 30 eyes had a central cRORA, one a paracentral cRORA, one an outer retinal atrophy, and three a macular neovascularization (MNV). At the second follow up, we found one more central and one more paracentral cRORA as well as one more MNV.

**Table 1.** patient demographics and fate of RPE lesions. Percentages of Number of eyes at follow ups refer to that at baseline. Percentages of females, males, and AMD progression refer to the number of subjects at the respective visit. Percentages of gain and loss of lesions refer to the number at baseline.

| **Patient information** | **baseline** | **follow up 1** | **follow up 2** |
| --- | --- | --- | --- |
| **N (patients)** | 37 (100%) | 25 (68%) | 11 (30%) |
| **female** | 23 (62%) | 14 (56%) | 5 (45%) |
| **male** | 14 (38%) | 11 (44%) | 6 (55%) |
| **age (years, mean±SD)** | 74.2±7.0 | 74.0±6.3 | 70.8±4.5 |
| **N (eyes)** | 40 (100%) | 30 (30%) | 12 (30%) |
| **eyes progressed to MNV** |  | 3 (10%) | 4 (33%) |
| **eyes progressed to atrophy** |  | 6 (20%) | 8 (67%) |
| **RPE lesions** | **N** | **gain(+)/loss(-)** | **gain(+)/loss(-)** |
| **thickened atop PED** | 58 (100%) | +5 (9%)/-6 (10%) | +11 (19%)/-9 (16%) |
| **hyperpigmentation** | 38 (100%) | +2 (5%)/-4 (11%) | +7 (18%)/-4 (11%) |
| **HRF** | 126 (100%) | +15 (12%)/-15 (12%) | +28 (22%)/-15 (12%) |

Hyperpigmentation and HRF have strong FAF with long lifetimes whereas thickened RPE on top of PED shows diffuse hyperfluorescence with short lifetimes. A typical example is shown in Fig. 1. This patient was imaged at baseline and at two follow up visits 24 and 52 months later. The natural history of PED is exemplified in two OCT sections. Superior to the macula we see a PED with hyperpigmentation at baseline. Its growth over 24 months is associated with the appearance of HRF (Fig 1e) showing strong focal hyperfluorescence with long lifetimes (at white arrowheads). At month 52, the PED is partly collapsed and partly denuded of RPE. Atrophy is seen by the penetration of laser light into the choroid in OCT (Fig. 1f) and a hypofluorescence with long lifetimes in LSC (Figs. 1p and v). Interestingly, this incipient atrophy does not show hypofluorescence but already long lifetimes in SSC. Whereas we hardly see RPE attached to the still elevated BLamD (red arrowhead in Fig 1f), HRF persist in the retina (Fig. 1f) and can contribute to the long lifetime fluorescence. Inferior to the macula, we see also growth of PED, which becomes hyperautofluorescent at month 24 (Figs. 1l and o, black arrowhead). In contrast to hyperpigmentation and HRF, PED have short FAF lifetimes. At month 52, we found these PEDs collapsed leaving migrated RPE with long lifetimes again (black arrowheads in figs 1c, i, m, p, s, and v). Finally, at month 52 we see one very small HRF (purple arrowhead in figs. 1c, j, m, p, s, and v) which is hardly discernible in CFP. FLIO, however clearly shows that this lesion has a long FAF lifetime.

**Figure 1.**
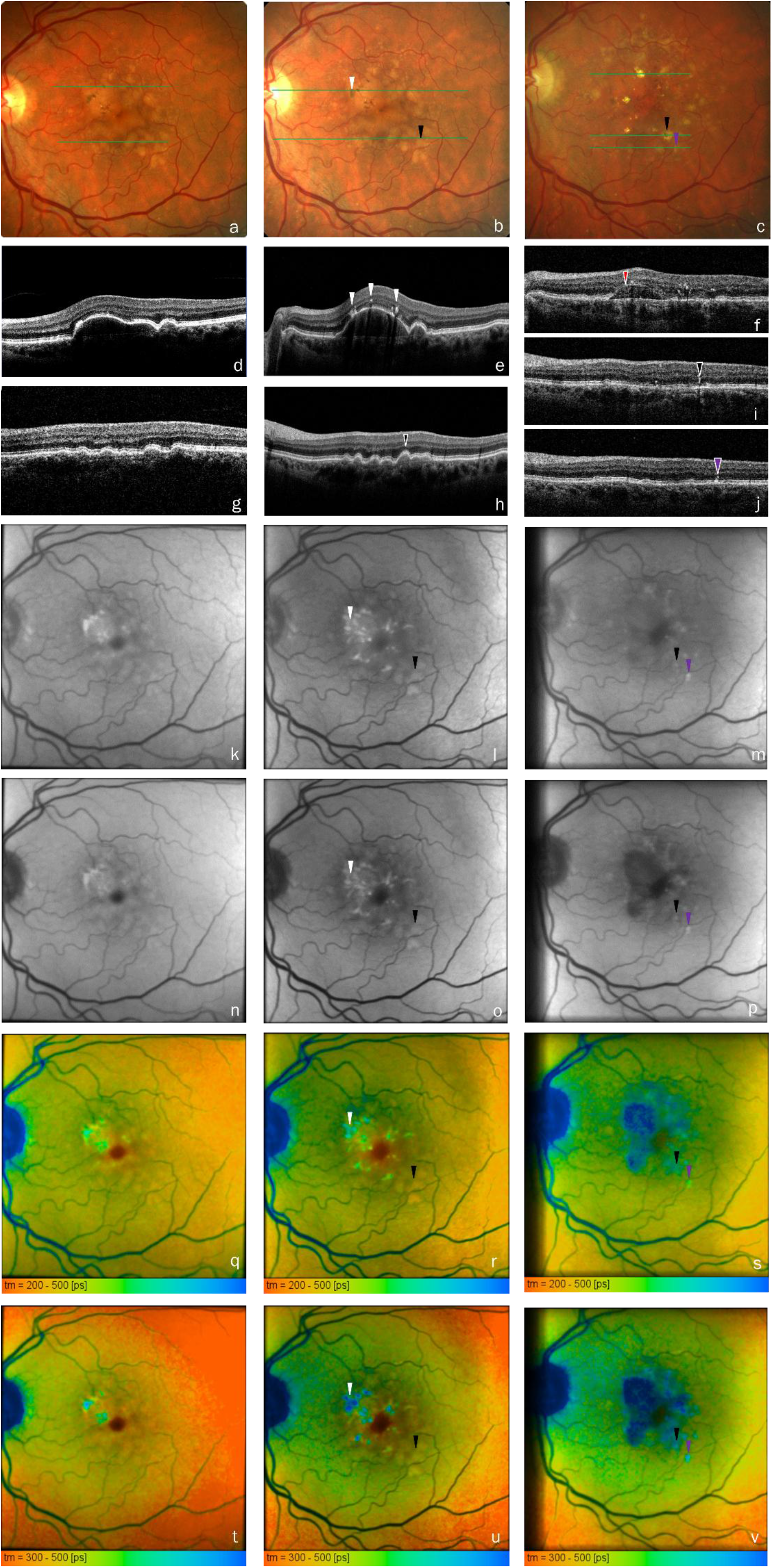
natural history of a PED with RPE migration. a-c: CFP, d – j: OCT, k – m: FAF SSC, n – p: FAF LSC, q – s: FLIO SSC, and t – v: FLIO LSC of a 64-year-old patient at baseline (left column: a, d, g, k, n, q, and t), 24 months (middle column: b, e, h, l, o, r, and u) and 52 months follow up (right column: c, f, I, j, m, p, s, and t). green lines in a – c show the localization of OCT scans. White arrowheads: HRF that correspond to hyperpigmentation on CFP. Red arrowhead: BLamD that persists after death or migration of RPE. Black arrowheads: growth of PED at months 24 leaving migrated RPE at month 52. Purple arrowhead: migrated RPE with HRF, no history of PED at this point, but clearly prolonged lifetimes in FLIO.

Whereas intraretinal HRF (i.e., internal to the ellipsoid zone) always show long lifetimes, in hyperpigmentation associated with focal thickening of the RPE-BL band we found a transition from short to long lifetimes. Fig, 2 shows an example (arrowheads point to hyperpigmented spots with various FAF lifetimes). At hyperpigmentation (Fig. 3, arrowhead), we observed a change from hyperfluorescence with initially short lifetime to long lifetime upon the appearance of HRF with no PED 35 months later. However, long lifetimes are also seen before the advent of atrophy in the absence of HRF. Fig. 4 shows a patient at baseline and 21 months later. The white arrowheads point to thickened RPE showing short lifetime, whereas long lifetimes also were found on PED (red arrowhead). Furthermore, HRF (black arrowhead), outer retinal atrophy (purple arrowhead), and cRORA (blue arrowhead) are seen.

**Figure 2.**
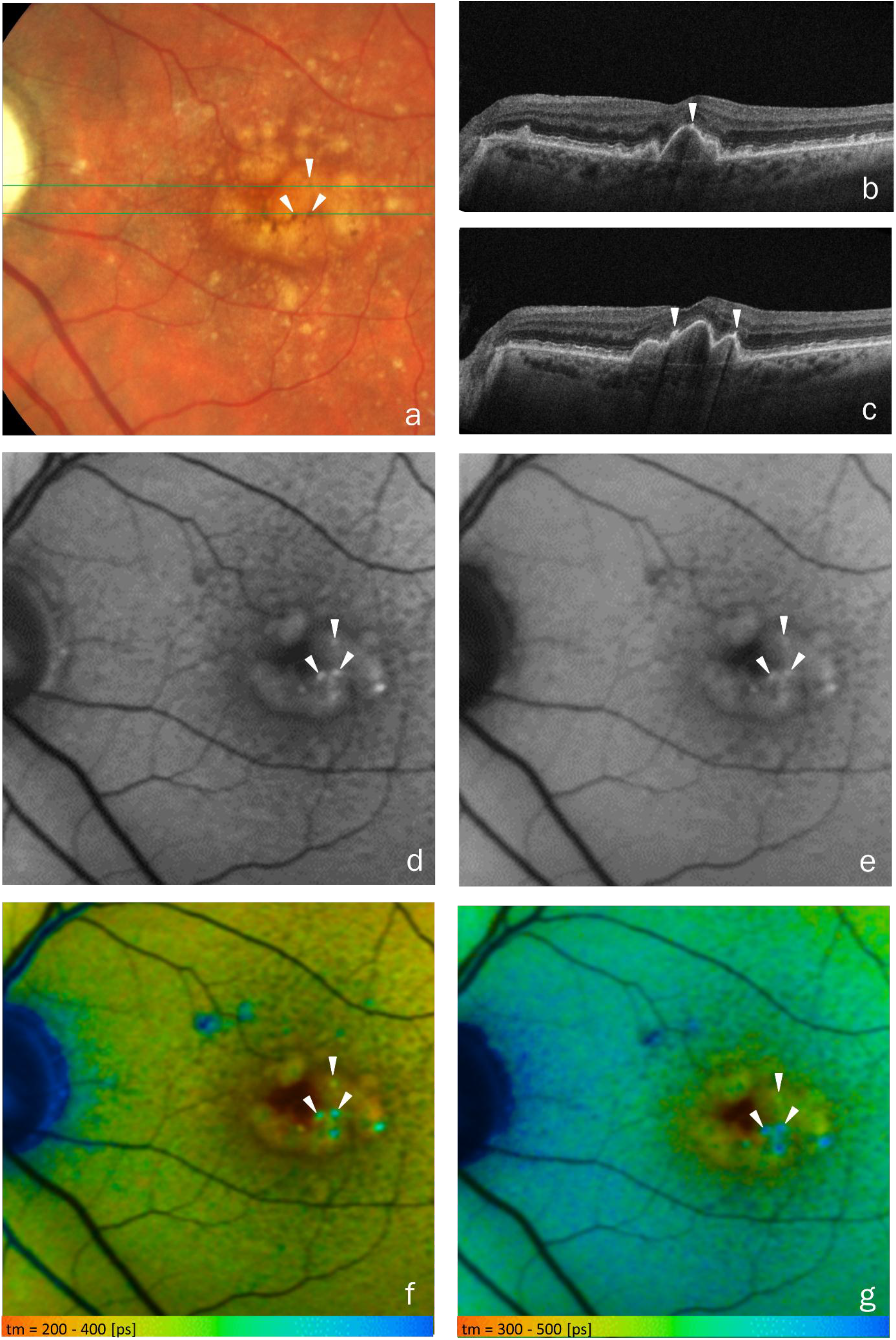
RPE detaching from the orthopic layer. a: CFP, b,c: OCT, d: FAF SSC, e: FAF LSC, f: FLIO SSC, and g: FLIO LSC of a 81-year-old patient. White arrowheads show corresponding hyperpigmentation (a), activated RPE cells (b-c), iso- and hyper-FAF (d,e), and lengthening lifetimes (f,g).

**Figure 3.**
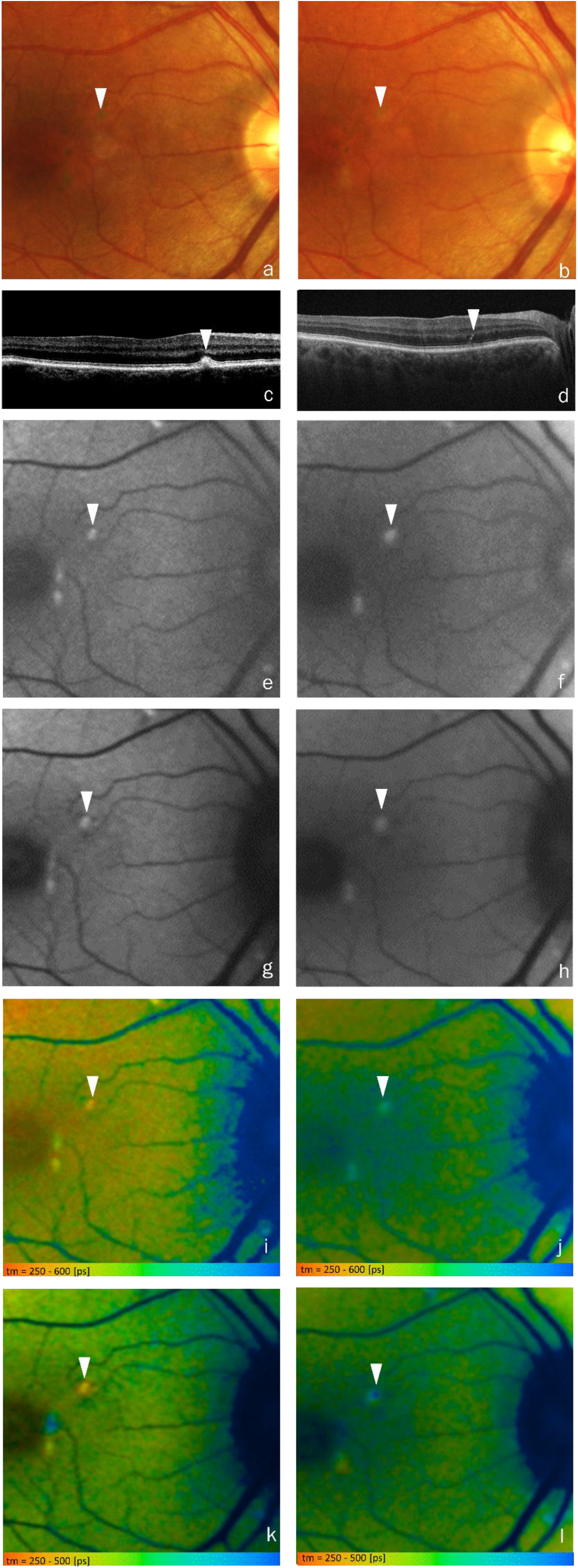
change of FAF lifetime of RPE in the conversion to HRF. a,b: CFP, c,d: OCT, e,f: FAF SSC, g,h: FAF LSC, i,j: FLIO SSC, and k,l: FLIO LSC of a 74-year-old patient at baseline (left column: a, c, e, g, i, and k) and 35 months follow up (right column: b, d, f, h, j, and l). The white arrowhead indicates a hyperpigmentation with short FAF lifetime atop a druse. At follow up, the druse was resorbed (no PED) and RPE changed to long FAF lifetime upon conversion to HRF. lt shows spatial heterogeneity in the changing of lifetimes.

**Figure 4.**
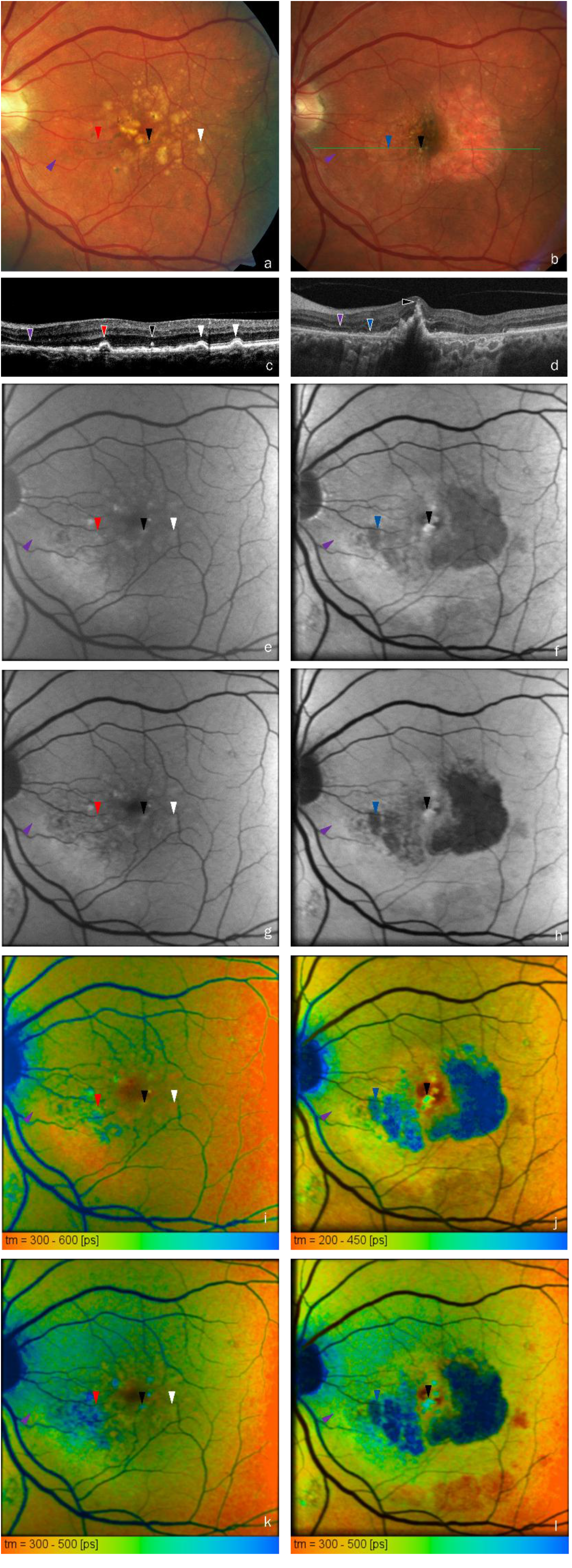
different lifetimes are found in activated and migrated RPE as well as outer retinal atrophy and cRORA. a,b: CFP, c,d: OCT, e,f: FAF SSC, g,h: FAF LSC, i,j: FLIO SSC, and k,l: FLIO LSC of a 69-year-old patient at baseline (left column: a, c, e, g, i, and k) and 21 months (right column: b, d, f, h, j, and l) follow up. Activated RPE atop PED (white arrowheads) shows short FAF lifetime, however also transition to long lifetime is seen (red arrowheads). Furthermore, migrated RPE (black arrowhead), outer retinal atrophy (purple arrowhead), and cRORA (blue arrowhead) are seen.

The average FAF lifetimes and ESIR, found for thickened RPE, hyperpigmentation as well as HRF, is given in Table 2 and shown as boxplots in Fig. 5. HRF have significantly longer lifetimes in both spectral channels and emit at shorter wavelengths (higher ESIR), than their respective environment. For hyperpigmentation, we found longer lifetimes in LSC only whereas they were significantly shorter in the thickened RPE atop PED than in the vicinity of these lesions in SSC. A comparison of the lesion types showed significantly longer FAF lifetimes for the hyperpigmentation and HRF than for thickened RPE (both spectral channels p<0.001). They were significantly longer for HRF than for hyperpigmentation in LSC (p<0.001); in SSC the difference was non-significant (p=0.374). The ESIR was significantly increasing from thickened RPE to hyperpigmentation to HRF, and the mean values were significantly different for all groups (p≤0.007). No correlation was found between the change of FAF lifetimes or ESIR of any lesion type and the time to follow up.

**Table 2.** RPE lesions at baseline. FAF lifetimes and ESIR (mean ± standard deviation) of RPE lesions at the lesion and in its vicinity, as well as the mean of individual differences (lesion – vicinity) of these measures with p-values from paired t-test.

|  |  | lesion | vicinity | mean difference | p for difference |
| --- | --- | --- | --- | --- | --- |
| thickened RPE-BL band | $\tau_m$ SSC | 256±49 ps | 261±56 ps | -5.0±9.0 ps | <0.001 |
| | $\tau_m$ LSC | 336±35 ps | 238±34 ps | -1.1±5.5 ps | 0.118 |
|  | ESIR | 0.552±0.079 | 0.547±0.088 | 0.005±0.022 | 0.087 |
| hyperpigmentation | $\tau_m$ SSC | 317±68 ps | 310±69 ps | 7.0±22.6 ps | 0.063 |
| | $\tau_m$ LSC | 377±56 ps | 265±50 ps | 11.9±15.5 ps | <0.001 |
|  | ESIR | 0.609±0.081 | 0.600±0.080 | 0.010±0.035 | 0.096 |
| HRF | $\tau_m$ SSC | 337±79 ps | 224±87 ps | 13.6±19.4 ps | <0.001 |
| | $t_m$ LSC | 414±50 ps | 389±46 ps | 24.8±18.1 ps | <0.001 |
|  | ESIR | 0.654±0.075 | 0.643±0.072 | 0.011±0.032 | <0.001 |

**Figure 5.**
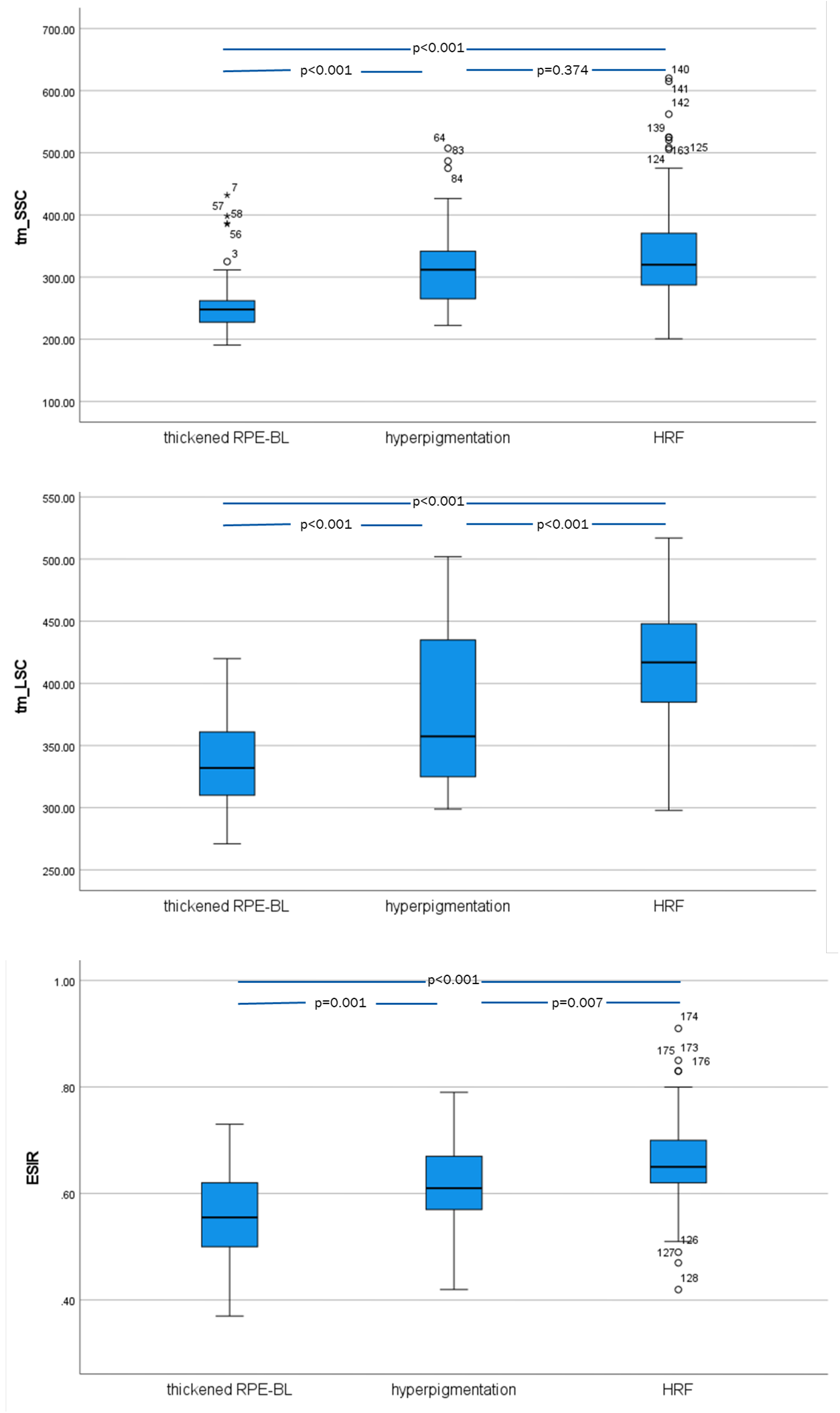
FAF lifetimes and ESIR. Boxplots of FAF lifetimes (top: SSC, middle: LSC) and ESIR (bottom) for activated RPE, hyperpigmentations, and migrated RPE. p-values are from post-hoc Bonferroni test after ANOVA.

## Discussion

RPE dysmorphia suggestive of pathologic activation is a key symptom of AMD. Shedding cells drop granule aggregates into the underlying basal laminar deposit (BLamD).^6^ Subducted cells are found between the RPE basal lamina and the inner collagenous layer of Bruch’s membrane,^44^ and acquired vitelliform lesions may result from the expulsion of organelles.^40, 45^ Some of these alterations can be seen in CFP as hyper- or hypopigmentation due to modulation of light-absorptive melanin and RPE thickening in OCT. Clinico-pathologic correlation revealed that HRF indicate single and clustered RPE cells that detached from their BL and migrated into the retina.^45^ Hyperpigmentation and RPE migration (HRF) are known indicators for the risk of AMD progression to cRORA.^4, 46^

The principal result of our study is the finding of a transition from short to long lifetimes, along with a hypsochromic shift of the emission spectrum, in the compromised RPE. Whereas hyperpigmentation shows longer lifetimes than its environment only in LSC, a switch to long lifetimes in both channels is seen after cells activate and detach from the RPE layer and migration beyond the ellipsoid zone. We could not completely elucidate, at which point in the transition of the RPE cells from activation to atrophy this switch happens. Figs. 2 – 4 indicate a gradual change of lifetimes not only for cells sloughed into the subretinal space, but also for in-layer cells still attached to Bruch’s membrane. Whether these cells were dissociated and what role BLamD^39^ plays if any remains to be determined by techniques providing higher resolution.

Furthermore, it is not clear yet, whether the focal prolongation of lifetimes at hyperpigmentation and HRF is associated to a general prolongation of lifetimes in AMD patients in an annulus corresponding to the outer ring of the Early Treatment of Diabetic Retinopathy Study grid (inner diameter 3 mm and outer diameter 6 mm, centered on the fovea).^30^

Hyperpigmentation as well as HRF show hyper-FAF. This could be explained by the fact that a monocellular RPE layer has been replaced by stacking or clumping of cells. However, the prolongation of FAF lifetimes as well as the shift of the emission spectrum indicate also a change in fluorophore composition or intracellular environment. An alternative explanation of the hyperautofluorescence of hyperpigmentation and HRF with long lifetimes would be fluorescence from collagen IV^29^ accumulated in BLamD.^39, 47^ However, HRF represent RPE cells migrated off their BL and the long lifetimes seem to be restricted to the spots of HRF. Thus, collagen IV in BLamD has to be questioned as a source of this particular long lifetime fluorescence.

In agreement with Sauer et al.,^48^ thickened RPE atop drusenoid PED showed increased FAF intensity with shortened lifetime in SSC. Although it can be questioned whether a statistically significant 5 ps shortening of FAF lifetime has a biological relevance, this might have various explanations: Firstly, if fluorophore composition is altered as well, the alteration differs from that in hyperpigmentation and HRF. Secondly, we might see fluorophores other than those in Lipofuscin and Melanolipofuscin, as their emission maximum is in the LSC,^49, 50^ but FAF lifetime shortening is seen in SSC. Thirdly, the additional fluorescence could originate from the drusenoid PED and not from the RPE. This, however, is questionable as hyperfluorescent drusen tend to have longer lifetimes.^51^ This was more clearly seen for drusen in areas of GA, i.e. not covered by RPE [Schultz et al., submitted to acta] as well as in histopathology.^52^

Taken together, FLIO, in combination with other imaging modalities, suggests the following natural history of disease progression from PED to atrophy: First RPE cells were pathologically activated resulting in thickening and hyperfluorescence. In this state, we see a growth of the PED, indicating a higher rate of basal deposition of material by the cells or an increased resistance for clearance through Bruch’s membrane and choriocapillaris^53, 54^ or both. Subsequently, hyperpigmentation is observed, which was often found to be associated to RPE cells sloughing from the layer and migration of the cells into the retina in histology.^5, 8, 10, 40, 45, 55-59^ As clinical imaging and histology revealed RPE migration towards retinal vessels^40^ as along Bruch’s membrane (subducted cells),^6^ we may speculate that deficiency in oxygen and/or nutrients, especially glucose,^60^ is a driving force for migration. Migration of some cells along with death of others that shed organelles into BLamD (resembling apoptosis) may be the reason for dissociation of cells remaining attached to Bruchs’s membrane. Finally, death of RPE together with that of the photoreceptors results in cRORA. Whereas dissociated and subducted cells are known from histology,^44, 61^ they were not seen in our investigation due to the limited resolution of OCT and FLIO.

Usually, FAF is associated to Lipofuscin and Melanolipofuscin. However, both contain mixtures of different compounds with different fluorescence properties. Eldred and Katz described ten fluorescent species and recorded their spectra.^49^ A change of the mix of different RPE granules over age is reported and Ach et al. hypothesize that Lipofuscin and Melanolipofuscin granules can undergo formation, maturation, and deletion in a dynamic process.^18^ Thus, a change of the fluorophore composition of RPE, either degenerating or rescuing itself by migration, is not unlikely. Different phenotypes of RPE granules, associated with hypo- or hyperfluorescence of the cells, are described.^62,63^ This further hints at possible changes of FAF properties in the course of cellular atrophy, however, a direct comparison is difficult the prior authors^62, 63^ did not measure FAF lifetimes or spectra and the resolution in our investigation is insufficient to resolve single granules. High-resolution microscopy of RPE flat mounts revealed degranulation and granule aggregation in AMD donor eyes.^18^ According to the authors, this may explain the clinical observation of reduced FAF in aged eyes (above 70 years). The RPE alterations, found in our study, however, show hyperfluorescence. Thus, possibly long FAF lifetimes in FLIO indicate compromised RPE at an earlier stage in the pathway to atrophy. A grading of RPE changes relevant to autofluorescence was introduced by Rudolf et al.^25^ and extended by Zanzottera et al.^6^ Although initially intended to describe RPE changes at the rim of GA,^64^ it categorizes similar lesions as we find here, including non-uniformity of pigmentation and morphology, sloughing, intraretinal migration, and loss of RPE. Interestingly, these authors did not find alterations of the histologic autofluorescence in stage 1 (non-uniformity of pigmentation and morphology) despite an abnormal expression of CD46, MTC3, and ezrin, indicating complement activation and loss of cellular polarity.^7^ This, again, suggests that fluorescence intensity is less sensitive to early RPE alterations than lifetime.

Along with the change of fluorescence lifetime and an emission spectrum shift, cells may alter their properties and function.^65^ A current immunohistochemistry investigation showed that abnormal RPE lose typical markers like the retinoid isomerohydrolase RPE65. On the other hand, they may gain properties of immune cells as shown by immunoreactivity for macrophage markers CD68 and CD163,^66, 41^ Thus, HRF have been considered macrophages or microglia that phagocytosed RPE.^67, 68^ However, HRF can survive for years in the retina and the distribution of granules was found to be preserved in histology.^6^ Therefore, a transdifferentiation of the RPE cells has to be considered.^5^ This transdifferentiation may be associated to the switch of fluorescence properties, however, the mechanism as well as contributing fluorophores still have to be determined.

Although FLIO primarily measures FAF lifetimes, the use of two spectral channels also enables a rough estimation of spectral FAF emission characteristics. This links RPE pathology to hypsochromic shift. Similarly, Marmorstein et al. found a shorter emission maximum for AMD than for control RPE in histology, however for an excitation wavelength of 364 nm.^69^ Three different spectra were revealed for lipofuscin and melanolipofuscin.^70^ Hyperspectral FAF imaging in RPE/choroid flatmounts of AMD donor eyes found the same spectra,^19^ however it remains to be determined, whether their abundance changes with RPE pathology as our data suggest. The existence of different fluorophore components, which can be distinguished by their spectra, supports our hypothesis that the fluorophore composition changes in the process of RPE activation and migration.

The following limitations of the current investigation have to be mentioned: This is a retrospective study of clinic patients. Thus, only some patients have follow up data, and the follow up intervals were not uniform. That limits our ability to draw conclusions on the progression timeline. This retrospective study includes also patients with varying quality of OCT. Therefore, a number of RPE lesions could not be classified unambiguously, and they were excluded from the analysis. Finally, as a non-invasive clinical imaging technique, FLIO cannot elucidate the chemical nature of the fluorophores exhibiting altered fluorescence lifetimes and emission spectra seen in this study.

In conclusion, we showed that thickening of RPE, hyperpigmentation, and formation of HRF is associated with specific changes in FAF, linking FAF to known progression indicators. Whereas initial activation of cells atop drusenoid PED is associated with a shortening of FAF lifetimes in SSC, a switch to long lifetimes was seen after cells sloughed from the intact layer or migrated anteriorly. Thus, FLIO might be used as an early indicator of RPE changes finally leading to atrophy. It should be used in conjunction with OCT to determine the health condition of RPE and can be used to study cellular changes which might indicate a risk for AMD progression.

## Data Availability

All data is available at the authors.

